# Beyond Pain Control: Exploring Regional Anaesthesia’s Potential to Improve Gastric Cancer Surgery Survival Rates: A Retrospective Cohort Study

**DOI:** 10.1101/2024.01.12.24301216

**Authors:** Onishi Tatsuki, Yoshika Onishi

## Abstract

**Background:** Gastric cancer management remains fraught with high morbidity and mortality rates owing to surgical interventions. Postoperative complications and extended hospitalisation profoundly affect patient survival, with elderly and medically compromised patients facing greater risks. We hypothesised that regional anaesthesia may reduce the surgical stress response and thus mortality. Hence, this study was aimed at investigating whether adjunctive application of regional anaesthesia during gastric cancer surgery attenuates the stress response to surgery, thereby improving postoperative survival outcomes.

**Methods:** In this study, we critically examined the potential role of regional anaesthesia in modulating perioperative immune and inflammatory responses, which are vital for improving oncological outcomes. Previous studies using diverse methodologies have provided heterogeneous and inconclusive results regarding the effectiveness of regional anaesthesia in enhancing cancer-related survival. Our study addresses these inconsistencies using a robust research design that was focused on a well-defined patient cohort.

**Results:** Preliminary evidence suggests that regional anaesthesia can modulate immune responses. However, its clinical benefits in cancer recurrence and survival have not been consistently demonstrated. The extant literature reflects a gap between preclinical promise and clinical efficacy, with no substantial evidence supporting a reduction in cancer recurrence after gastroesophageal cancer surgery under regional anaesthesia.

**Conclusion:** This study provides new insights into the role of regional anaesthesia in reducing the overall mortality in gastric cancer surgery. By employing stringent inclusion criteria and a well-defined patient cohort, this study aimed to provide clarity in a field marked by inconclusive evidence and guide future clinical practices and high-quality research initiatives.

## Introduction

Management of postoperative complications in elderly patients with gastric cancer is crucial because these complications are strongly affect clinical outcomes.^1^ Age, comorbidities, extent of resection, operation time, and combined resection are predictive factors for surgical complications in elderly patients.^2^ Factors such as age, splenectomy, and extended lymphadenectomy are associated with the development of complications after gastric cancer surgery.^3^ Despite these challenges, age is not a contraindication for the curative resection for gastric cancer, and surgical strategies should be modulated based on comorbidities, tumour stage, and future quality of life.^4^ Advances in aspects related to pre-, intra-, and postoperative stages of gastric cancer surgery, including preoperative risk assessment, fast-track programs, laparoscopic surgery, and optimal timing of oral diet, have the potential to improve outcomes.^5^ The safety and excellent prognosis associated with gastrectomy in elderly patients have been demonstrated, suggesting that it can be very safely performed by specialists.^6^

The use of regional anaesthesia in gastric cancer surgery is a topic of ongoing research with potential implications for patient outcomes. Studies have shown that regional anaesthesia, particularly thoracic epidural analgesia (TEA), can attenuate the stress response to surgery and facilitate gastrointestinal recovery.^7^ However, the effect of regional anaesthesia on cancer outcomes remains unclear.^8^ The use of multimodal analgesic strategies, including regional anaesthesia, has been associated with improved postoperative outcomes.^9^ Despite the potential benefits of regional anaesthesia, evidence is currently insufficient to warrant a change in clinical practice.^8^ Further research is required to determine the optimal anaesthesia technique for cancer surgery.^10^ A body of research suggests a potential link between regional anaesthesia and reduced mortality in cancer surgery, with particular focus on the impact of anaesthetic techniques and perioperative factors.^11^ In particular, amide-type local anaesthetics have shown promise in inhibiting cancer metastases.^12^ However, the evidence is inconclusive, with some studies indicating no significant differences in cancer recurrence rates with the use of regional anaesthesia.^13^ The potential benefits of regional anaesthesia in modulating immune and inflammatory responses during gastrointestinal cancer surgery have also been highlighted.^14^ Despite these findings, the impact of regional anaesthesia on cancer recurrence in patients with late-stage cancer remains controversial.^15^ Further research is required to establish a clear connection between regional anaesthesia and reduced mortality in cancer surgery.

The impact of regional anaesthesia on cancer-related outcomes remains inconclusive, with conflicting evidence from both preclinical and clinical studies.^14, 16^ While some studies have suggested a potential benefit in reducing cancer recurrence and improving survival,^17, 18^ others have found no significant association.^19-22^ The lack of standardized outcome definitions, low study quality, and high heterogeneity further complicate the interpretation of these findings.^14, 18^ Therefore, the use of regional anaesthesia in cancer surgery remains a topic in need of further high-quality research.^14, 20, 21^

While there is a body of work examining the impact of regional anaesthesia on cancer outcomes, the evidence remains inconclusive. Our research could potentially fill this gap by providing new insights into the role of regional anaesthesia in reducing overall mortality associated with gastric cancer surgery. We meticulously investigated the impact of concomitant regional anaesthesia on the overall mortality rates in patients undergoing gastric cancer surgery. We used a well-defined patient cohort with stringent inclusion criteria to ensure the relevance and precision of our findings.

## Methods

This study was approved by the Tokyo Metropolitan Bokutoh Hospital Ethics committee as 30-110 and 715 patients were retrospectively selected in accordance with the Declaration of Helsinki. The participants were categorized based on key demographic and clinical parameters including age and sex, allowing for a comprehensive analysis of the effects of regional anaesthesia across diverse patient subgroups. Rigorous statistical methods, including Kaplan-Meier analysis and Cox proportional hazards modelling, were applied to evaluate mortality outcomes.

**Table 1:**
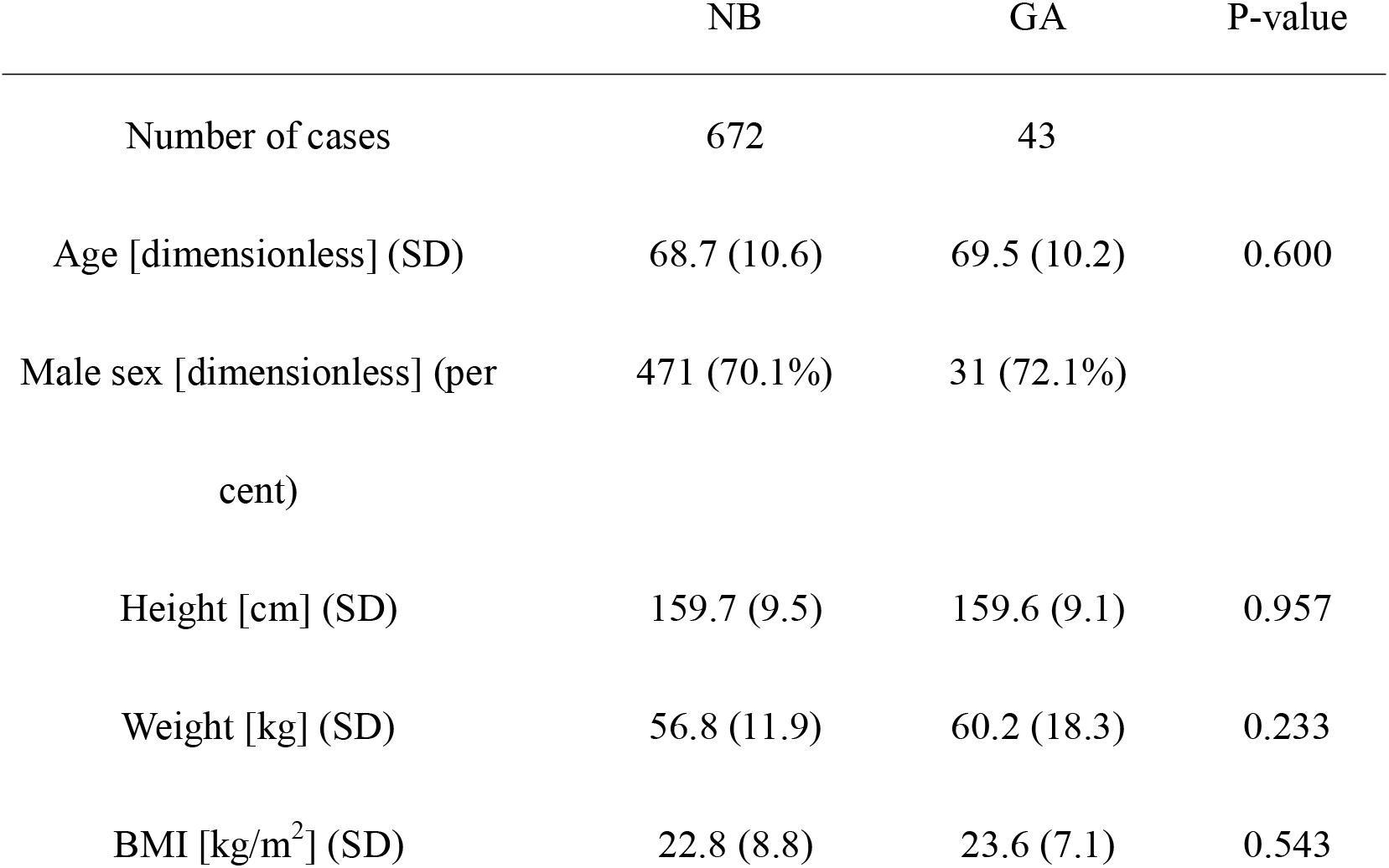
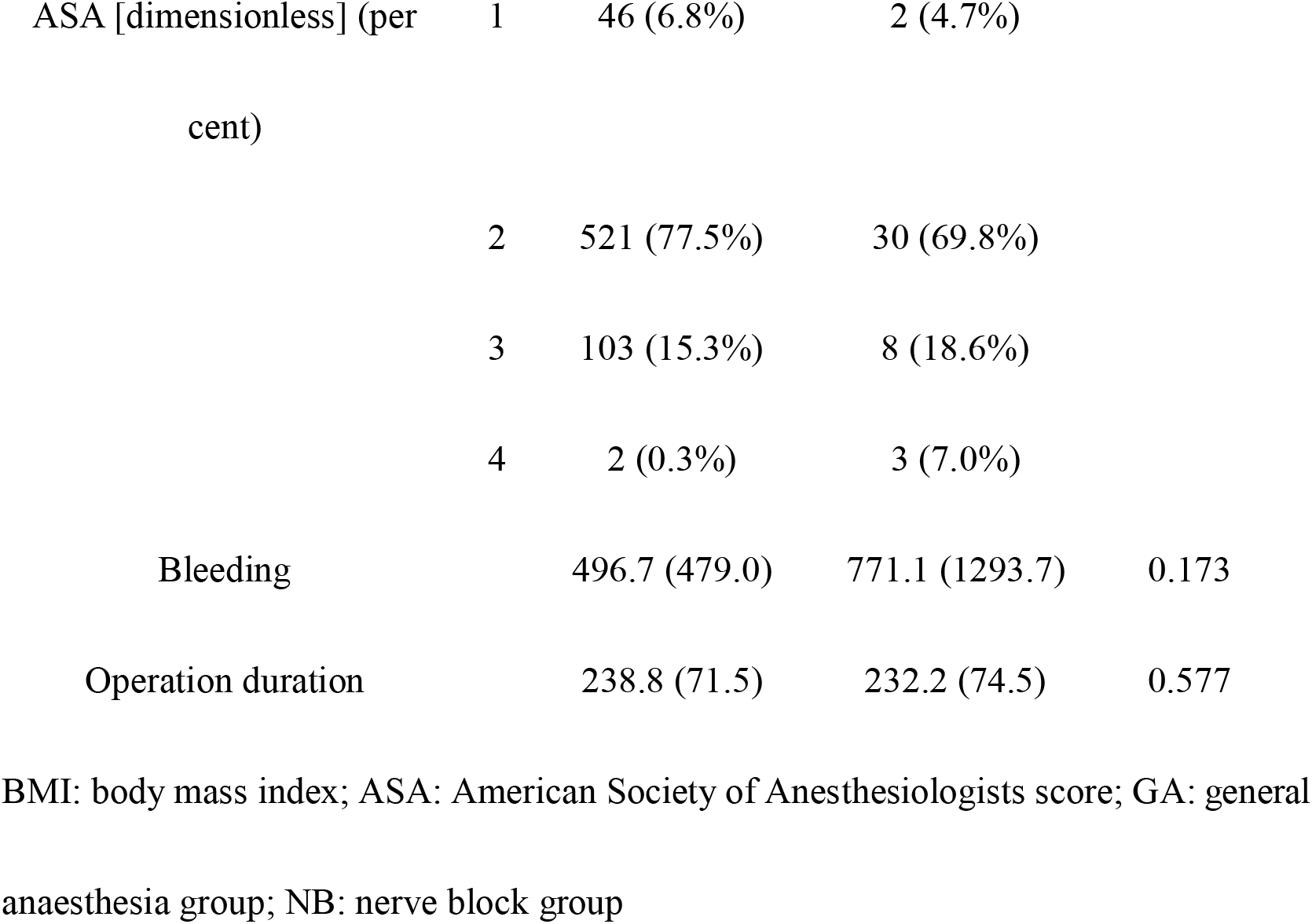
Characteristics and demographics of cases.

|  | NB | GA | P-value |
| --- | --- | --- | --- |
| Number of cases | 672 | 43 |  |
| Age [dimensionless] (SD) | 68.7 (10.6) | 69.5 (10.2) | 0.600 |
| Male sex [dimensionless] (per cent) | 471 (70.1%) | 31 (72.1%) |  |
| Height [cm] (SD) | 159.7 (9.5) | 159.6 (9.1) | 0.957 |
| Weight [kg] (SD) | 56.8 (11.9) | 60.2 (18.3) | 0.233 |
| BMI [kg/m<sup>2</sup>] (SD) | 22.8 (8.8) | 23.6 (7.1) | 0.543 |

|  |  |  |  |  |
| --- | --- | --- | --- | --- |
| ASA [dimensionless] (per cent) | 1 | 46 (6.8%) | 2 (4.7%) |  |
|  | 2 | 521 (77.5%) | 30 (69.8%) |  |
|  | 3 | 103 (15.3%) | 8 (18.6%) |  |
|  | 4 | 2 (0.3%) | 3 (7.0%) |  |
| Bleeding |  | 496.7 (479.0) | 771.1 (1293.7) | 0.173 |
| Operation duration |  | 238.8 (71.5) | 232.2 (74.5) | 0.577 |

BMI: body mass index; ASA: American Society of Anesthesiologists score; GA: general anaesthesia group; NB: nerve block group

## Results

The cumulative incidence curves allowed a comparison of the effects of concomitant nerve blocks on death over time (Fig. 1). The graph indicated that all-cause mortality was higher in the group administered general anaesthesia (GA) alone than in the combined nerve block (NB) group. Specifically, at 5000 days of follow-up, approximately 80% of the patients in the GA group died, compared with approximately 30% in the NB group. This result suggests that interventions or conditions in the GA group may increase the risk of all-cause mortality compared with those in the NB group.

**Figure 1:**
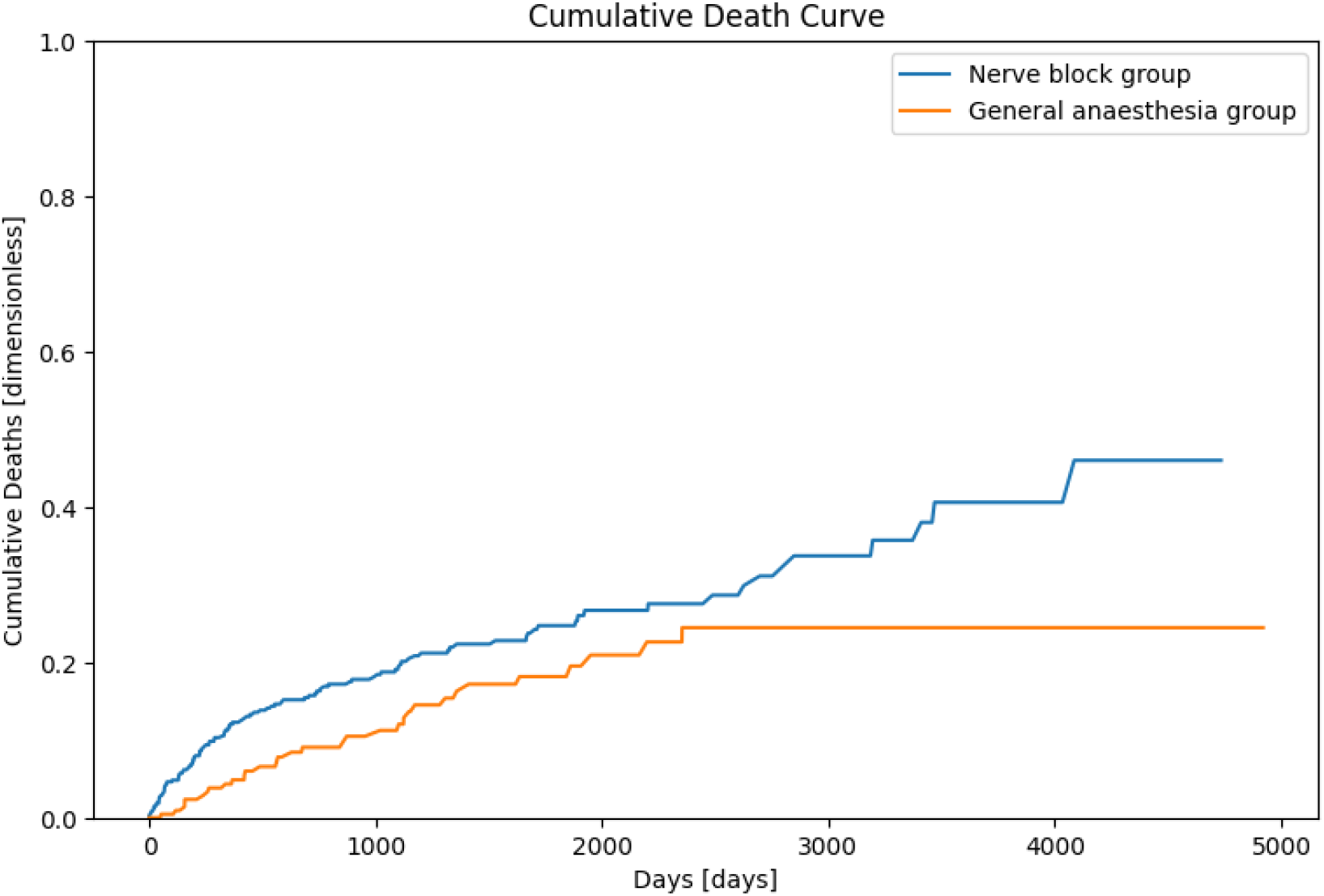
Cumulative deaths.

## Discussion

Management of postoperative complications in patients with gastric cancer is strongly associated with clinical outcomes and involves predictors such as age, complications, extent of resection, operative time, and resection.^1-3^ Advances in each stage of gastric cancer surgery, including preoperative risk assessment, fast-track programs, laparoscopic surgery, and optimal timing of oral diet, have the potential to improve outcomes.^5^ Specialists have demonstrated the safety and excellent prognosis of gastrectomy.^6^

The use of regional anaesthesia may influence the outcomes of gastric cancer surgery, particularly thoracic epidural analgesia (TEA), which attenuates the stress response to surgery and facilitates gastrointestinal recovery.^7^ However, the effects of regional anaesthesia on cancer outcomes remain unknown.^8^ The use of multimodal analgesia strategies has been associated with improved postoperative outcomes; however, there is insufficient evidence to warrant a change in clinical practice.^9, 10^

An association between regional anaesthesia and lower mortality in cancer surgery has been suggested, with amide local anaesthetics in particular showing promise in reducing cancer metastasis.^11, 12^ However, some studies found no significant differences in cancer recurrence rates with the use of regional anaesthesia,^13^ and the potential benefits of regional anaesthesia in modulating immune and inflammatory responses during gastrointestinal cancer surgery remain controversial.^14, 15^ The impact of regional anaesthesia on cancer-related outcomes is inconclusive, with conflicting evidence from preclinical and clinical trials.^14, 16-22^ The lack of standardised outcome definitions, low study quality, and high heterogeneity complicate the interpretation of these findings. Regional anaesthesia may improve overall survival, but does not necessarily reduce cancer recurrence.^14, 23-25^ A variety of findings have shown that general intravenous anaesthesia with propofol improves survival associated with gastric cancer surgery; however, there is no significant association between epidural anaesthesia and long-term survival.^26-28^

This study examined how the combined use of regional anaesthesia affects the survival of patients with gastric cancer. On the basis of our theoretical framework, we hypothesised that regional anaesthesia modulates the neuroendocrine stress response to surgery, thereby reducing systemic inflammation and improving survival outcomes. The present study demonstrated that the concomitant use of regional anaesthesia in gastric cancer surgery may improve survival outcomes.

## Data Availability

All data produced in the present study are available upon reasonable request to the authors

## Author contributions

O.T.: concept, initial draft, data processing, visualization, final approval Y.O.: concept, final approval

## Acknowledgements

Tatsuyoshi Ikenoue at Shiga University, Data Science and AI Innovation Research Promotion Center

## Code availability

Data should be available upon request. Codes were uploaded at https://github.com/bougtoir/gastric_nb_death

## Funding

